# Prolonged presence of SARS-CoV-2 in feces of pediatric patients during the convalescent phase

**DOI:** 10.1101/2020.03.11.20033159

**Authors:** Yuhan Xing, Wei Ni, Qin Wu, Wenjie Li, Guoju Li, Jianning Tong, Xiufeng Song, Quansheng Xing

## Abstract

**Background:** Severe acute respiratory coronavirus 2 (SARS-CoV-2) is a newly identified virus which mainly spreads from person-to-person. Presence of SARS-CoV-2 has been constantly reported in stools of patients with coronavirus disease 2019 (COVID-19). However, there is a paucity of data concerning fecal shedding of the virus in pediatric patients.

**Objective:** To investigate dynamic changes of SARS-CoV-2 in respiratory and fecal specimens in children with COVID-19.

**Methods:** From January 17, 2020 to February 23, 2020, three pediatric cases of COVID-19 were reported in Qingdao, Shandong Province, China. Epidemiological, clinical, laboratory, and radiological characteristics and treatment data of these children were collected. Real-time fluorescence reverse-transcriptase-polymerase-chain reaction (RT-PCR) was performed to detect SARS-CoV-2 RNA in throat swabs and fecal specimens. Patients were followed up to March 10, 2020, the final date of follow-up, and dynamic profiles of RT-PCR results were closely monitored.

**Results:** All the three pediatric cases were household contacts of adults whose symptoms developed earlier. Severity of disease was mild to moderate and fever was the most consistent and predominant symptom at onset of illness of these children (two cases had body temperature higher than 38.5°C). All children showed increased lymphocytes (>4.4×10^9^ /L) with normal white blood cell counts on admission. Radiological changes were not typical for COVID-19. All children showed good response to supportive treatment. Clearance of SARS-CoV-2 in respiratory tract occurred within two weeks after abatement of fever, whereas viral RNA remained positive in stools of pediatric patients for longer than 4 weeks. Two children had fecal SARS-CoV-2 turned negative 20 days after throat swabs showing negative, while that of another child lagged behind for 8 days.

**Interpretation:** SARS-CoV-2 may exist in gastrointestinal tract for a longer time than respiratory system. Persistent shedding of SARS-CoV-2 in stools of infected children indicates the potential for the virus to be transmitted through fecal excretion. Massive efforts should be made at all levels to prevent spreading of the infection among children after reopening of kindergartens and schools.

## Introduction

On December 12, 2019, 27 pneumonia cases of unknown cause emerged in Wuhan, Hubei Province, China.^1^ The etiological agent was identified as a novel coronavirus and was later renamed as severe acute respiratory coronavirus 2 (SARS-CoV-2) by the world health organization (WHO).^2,3^ The pandemic of coronavirus disease 2019 (COVID-19) has wreaked havoc in China and spread rapidly to about ninety countries, constituting a global threat with the highest risk impact.^4^ Epidemiological evidence gained in China suggested that most individuals lack relevant immunity and are generally susceptible to the virus. The majority of published studies of COVID-19 focused on adult populations.^5-8^ While knowledge on SARS-CoV-2 infection in children are still yet to be fully developed and only a limited number of case reports of pediatric patients are currently available.^9-12^

As of February 23, 2020, a cumulative total of 60 confirmed COVID-19 cases were reported in Qingdao, Shandong Province, China, of which three cases (5%) were children under 10 years of age. Pediatric patients presented clinical characteristics distinct from those observed in adult patients. Notably, SARS-CoV-2 RNA was detected in the stool of these children 8-20 days after negative conversion of viral RNA in respiratory specimens. Whereas the majority of adult patients had negative results of nucleic acid testing in respiratory and fecal specimens synchronously. Prolonged presence of SARS-CoV-2 RNA in feces after showing negative in respiratory specimens may become a source of COVID-19 in the community and pose a threat to public health if fitness for discharge is based on the current version of *Diagnosis and Treatment Plan of Corona Virus Disease 2019* (“with normal body temperature for more than 3 days”, “with obvious features of absorption of inflammation shown in lung imaging” and “negative results of the nucleic acid tests of respiratory pathogens for consecutive two times [sampling interval at least 1 day]”).^13^

## Methods

### Patients

From January 17, 2020 to February 23, 2020, a total of 60 patients were diagnosed with COVID-19 in Qingdao, Shandong Province, China. We recruited all three pediatric patients with laboratory confirmed SARS-CoV-2 infection who were reported by the local health authority. Diagnosis of COVID-19 was based on the WHO interim guidance.^14^ Patients were followed up on a regular basis after hospital discharge till March 10, 2020.

This study was approved by the Ethics Commission of Qingdao Women and Children’s Hospital (QFFLL-KY-2020-11) and written informed consent was obtained from patients’ legal guardians prior to enrolment.

### Data collection

Personal, clinical, laboratory, and radiological characteristics and treatment and outcomes information were obtained with standardized data collection forms from electronic medical records. Additionally, we directly contacted patients’ families to ascertain epidemiological and symptom information. Data were entered into a computerized database and double-checked by two researchers independently.

### Sample Collection and Detection of SARS-CoV-2

Throat swabs were obtained from patients on admission. Fecal specimens were first collected in two patients (case 1, 1.5-year-old male; and case 2, 5-year-old male) on day 4 after onset of the disease. While the stool sample from another patient (case 3, 6-year-old female) was obtained 9 days after hospital discharge. To monitor the dynamic changes of viral shedding, we obtained throat swabs from patients every day during hospitalization and every other day during follow-up after discharge. Fecal specimens were collected applying a similar rationale since the first day of sample collection (samples from case 3 were only collected during follow-up period). Presence of SARS-CoV-2 RNA was detected by real-time fluorescence reverse-transcriptase-polymerase-chain reaction (RT-PCR) using a commercial kit approved by the China Food and Drug Administration. Detailed procedures were described elsewhere.^15^

## Results

### Clinical presentations, laboratory and radiological findings

Severity of disease in all patients was mild according to *Diagnosis and Treatment Plan of Corona Virus Disease 2019* (tentative fifth/sixth edition).^16,17^ The disease started with fever (≧38.5°C) in all of the three children. None of them developed severe complications nor required intensive care or mechanical ventilation. All children showed good response to supportive treatment including inhalation of interferon, oral Ribavirin and traditional Chinese medicine.

Laboratory findings of these children on admission to hospital are shown in **Table 1**. On admission, all the children had increased lymphocyte count (>4.4×10^9^ /L). Only case 1 showed decreased neutrophil count (<1.7×10^9^ /L), whereas those of the other two children were within normal range. Elevation of platelets was observed in case 1 and case 2. Case 3 had increased levels of procalcitonin (0.73 ng/mL) and C-reaction protein (10.5 mg/L), while serum level of D-dimer (860.0 ng/mL) was found elevated in case 1. Reduced level of serum creatinine was detected in case 1 and case 2 (<41.0 µmol/L). Transverse chest computed tomograms (CT) showed delicate patches of ground glass opacity of lower lobe of right lung in case 1 on day 1 after symptom onset and consolidation changes of the left lower lobe near the pleura in case 2 on admission; while case 3 had no abnormality on CT imaging (**Figure 1**).

**Table 1.** Laboratory Findings of Pediatric Patients with COVID-19 on Admission to Hospital

| Variables | Case 1 | Case 2 | Case 3 |
| --- | --- | --- | --- |
| White blood cell count ($\times 10^9/L$; normal range 4.0-12.0) | 7.3 | 9.6 | 6.0 |
| Neutrophil count ($\times 10^9/L$; normal range 1.7-7.7) | **1.2** ↓ | 3.6 | 1.7 |
| Lymphocyte count ($\times 10^9/L$; normal range 0.4-4.4) | **5.4** ↑ | **5.2** ↑ | **4.9** ↑ |
| Platelet count ($\times 10^9/L$; normal range 100.0-300.0) | **333.0** ↑ | **411.0** ↑ | 186.0 |
| Hemoglobin (g/L; normal range 310.0-370.0) | 332.0 | 359.0 | 332.0 |
| Alkaline phosphatase (U/L; normal range 0.0-500.0) | 179.0 | NA | **639.0** ↑ |
| Lactate dehydrogenase (U/L; normal range 120.0-250.0) | **264.3** ↑ | NA | 194.0 |
| Serum creatinine ($\mu\text{mol/L}$; normal range 41.0-73.0) | **22.8** ↓ | **28.3** ↓ | 53.4 |
| Creatine kinase (U/L; normal range 50.0-310.0) | 73.2 | 88.6 | 91.0 |
| Procalcitonin (ng/mL; normal range 0-0.5) | 0.23 | 0.21 | **0.73** ↑ |
| Erythrocyte sedimentation rate (mm/h; normal range 0-15.0) | 3.0 | 10.0 | 7.8 |
| C-reactive protein (mg/L; normal range 0-8.0) | <0.8 | <5.0 | **10.5** ↑ |
| D-dimer (ng/mL; normal range 0-550.0) | **860.0** ↑ | 230.0 | 190.0 |

Values out of normal range are shown in bold; ↑ denotes levels above the 99th percentile upper reference limit; ↓ denotes levels below the 1st percentile lower reference limit; NA= not available.

**Figure 1.**
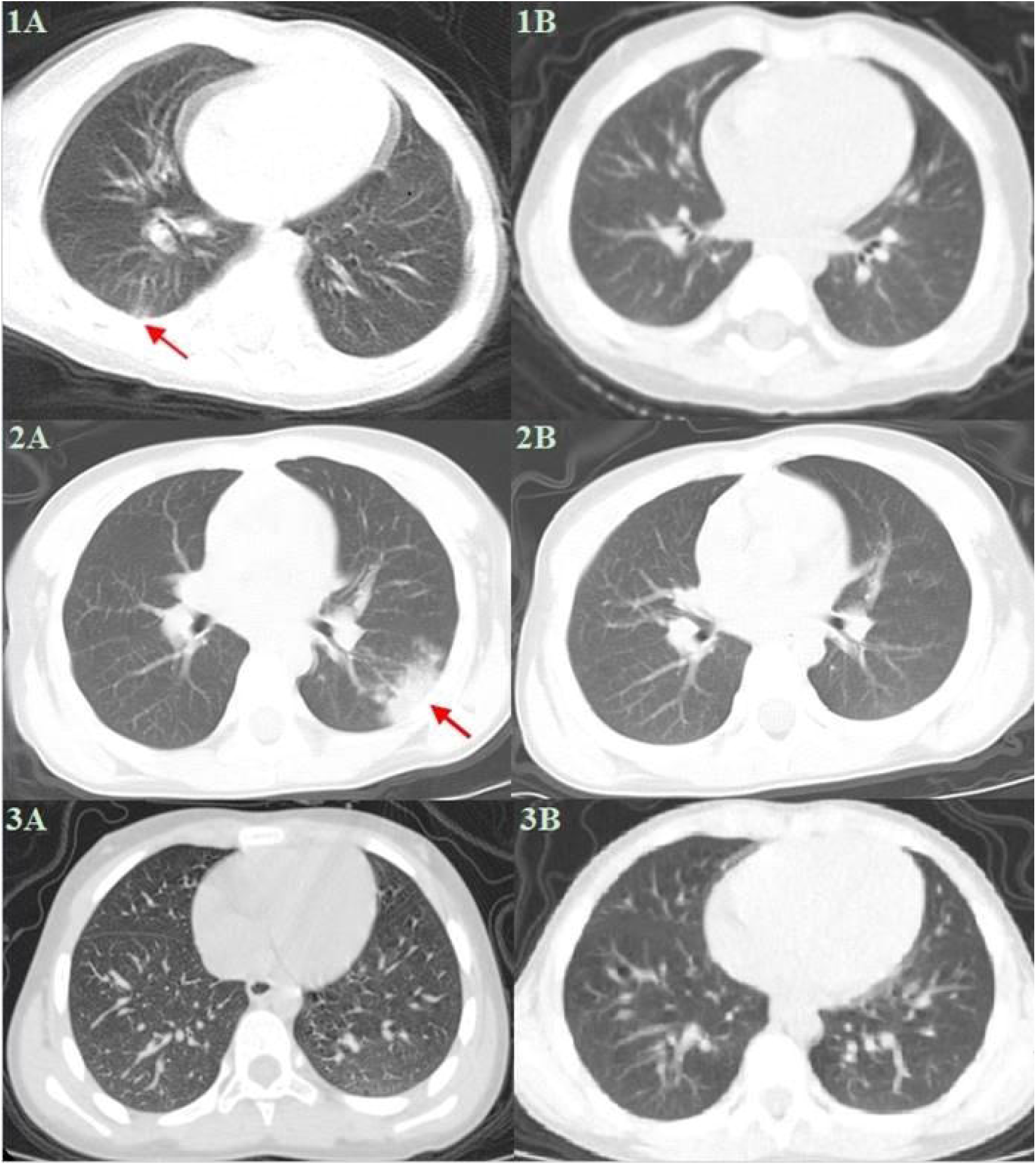
Transverse Chest Computed Tomographic Images. CT image from case 1 on day 1 after symptom onset, showing delicate patches of ground glass opacity of lower lobe of right lung near the pleura (1A, red arrow). After receiving treatment for 10 days, the lesions were completely absorbed (1B). Case 2 showed ground glass consolidation of the left lower lobe near the pleura on admission (2A, red arrow). Complete resolution of lesion was observed after treatment for 13 days (2B). There was no obvious abnormality in bilateral lung fields of case 3 on admission (3A). No change in CT imaging was obsetved during patient’s hospitalization (3B).

### Persistent fecal shedding of SARS-CoV-2 in pediatric patients during the convalescent phase

Two pediatric patients (case 1 and case 2) and their family members with SARS-CoV-2 infection were admitted in Qingdao Women and Children’s Hospital. During clinical practice, we noticed that the time for viral RNA in respiratory specimens turning negative was similar between pediatric patients and infected adults (these children’s family members). At this point, adult patients had negative results for nucleic acid testing in fecal specimens, whereas SARS-CoV-2 RNA remained detectable in stools from the two infected children. Therefore, we conducted a quarantine and surveillance protocol for the children and their family members who were already qualified for hospital discharge according to current standards after recovery with two consecutively negative RT-PCR test results in throat swabs (sampling interval at least 1 day). Concerned about the possibility of fecal-oral transmission, we recorded the timeline of changes in nucleic acid testing results in both throat swabs and fecal samples collected from these patients (**Figure 2**). All adult patients of the two families showed negative results for fecal SARS-CoV-2 RNA detection. Strikingly, viral RNA remained detectable in stools of the two children for 8 and 20 days, respectively, after nucleic acid turning negative in respiratory samples.

**Figure 2.**
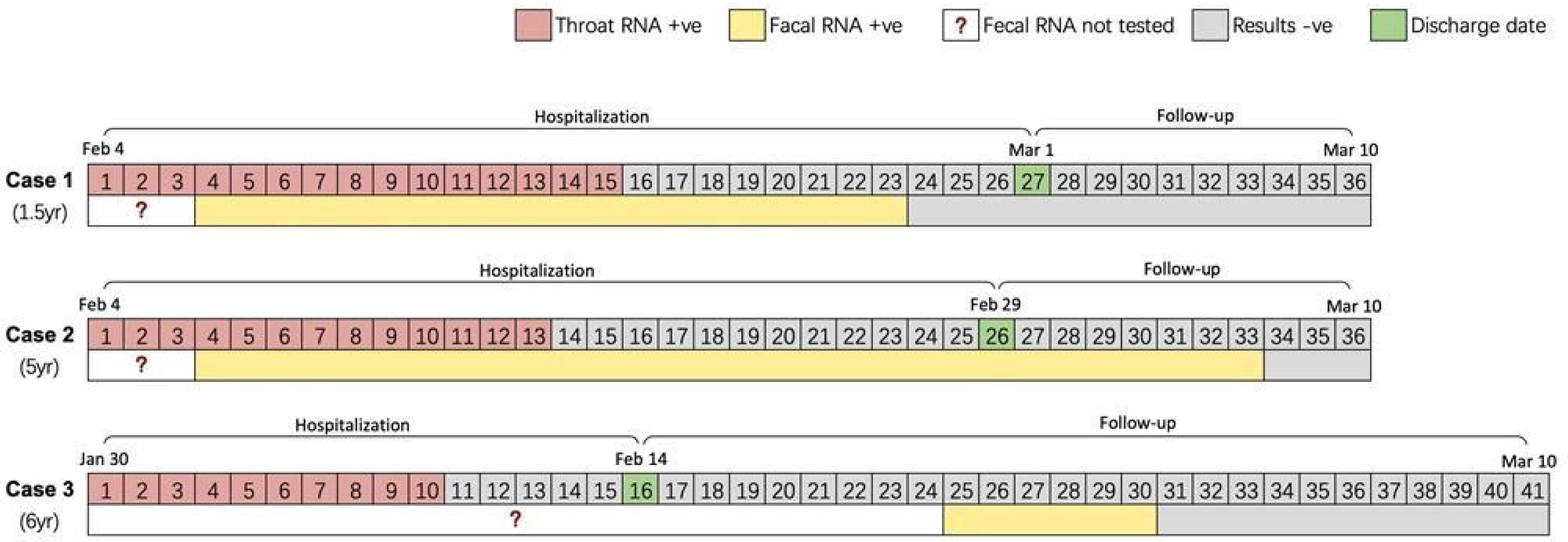
Dynamic Profile of SARS-CoV-2 RNA in Respiratory and Fecal Specimens. Days from admission are shown in the first row of each patient. Days filled in green represent the date of hospital discharge. Boxes filled in red denote the days which throat swabs were positive for RT-PCR analysis. Bars filled in yellow denote fecal specimens were consecutively positive for RT-PCR analysis. Boxes and bars filled in grey are the period which nucleic acid testing results were negative. The bars with an internal red question mark indicate days when fecal specimens were not collected and thus virological data were not available.

We suspected whether this phenomenon also existed in the third pediatric patient (case 3) who was admitted in another designated hospital in Qingdao. Unfortunately, by the time we reached the patient, this child had already been “cured” and discharged. Medical professionals of this hospital did not collect fecal specimens from the patient and they only performed RT-PCR testing for throat swabs plus CT according to *Diagnosis and Treatment Plan of Corona Virus Disease 2019* (tentative fifth edition).^16^ Municipal Centre of Disease Control and Prevention of Qingdao conducted nucleic acid testing in stool samples collected from case 3 and the child’s family members 9 days after hospital discharge. This child showed positive results for RT-PCR analysis in feces. During quarantine and follow-up period, clearance of SARS-CoV-2 in stool samples occurred 20 days after viral RNA in respiratory specimens turning negative. Detailed information was not available as no fecal sample was examined during hospitalization.

## Discussion

The newly issued report of the WHO-China Joint Mission on COVID-19 summarized the evidence so far on SARS-CoV-2 and pointed out 2.4% of those infected were individuals below 18 years of age.^18^ According to data released by the China Centers for Disease Control and Prevention, only 0.9% of COVID-19 patients were children under the age of 10 years.^19^ Among the 60 patients infected with SARS-CoV-2 in Qingdao, 3 (5%) cases were children younger than 10 years of age.

Most of the pediatric COVID-19 patients had mild disease with good treatment response and a relatively short time to resolution, with exception of one critically ill case reported in Wuhan.^12^ Consistent with previous findings, the three infected children in our study only presented with fever and mild cough or with no obvious symptom but CT abnormalities. From available data, children appeared to be slightly affected by SARS-CoV-2, a feature resembling that of SARS-CoV emerged 17 years ago.^20^ However, the relatively low attack rate of COVID-19 in children could be explained by the stringent implementation of home confinement and prolonged school closure during the outbreak as required by the Chinese governments. Whether it is also the case during school year is hard to tell. One distinct feature of pediatric cases is that almost all of them are infected through household contact with adult patients.^9-12,21^ As with all new diseases, many characteristics of COVID-19 still remain largely unknown. There is limited data to support the notion that children are less susceptible to SARS-CoV-2 infection or virus transmission is less effective among them.

Viral RNA has been constantly detected in stool samples and anal swabs collected from confirmed cases of COVID-19.^5,22-25^ In one study, fecal specimens from 9 (53%) of 17 patients were positive for nucleic acid testing, although viral loads of the stool were less than those of respiratory samples.^24^ Presence of SARS-CoV-2 was even detected in environmental samples taken from the surface of toilet bowl and sink in infection isolation rooms.^26^ Moreover, SARS-CoV-2 remained viable in the stool of infected patients as reported by some case reports.^18^ Evidence so far indicates the potential for SARS-CoV-2 to be transmitted through fecal excretion.^18^ Although the role of fecal shedding in viral transmission has not been systematically determined, a cautious approach is warranted when handling stool samples.

While we were closely monitoring the dynamic changes of fecal SARS-CoV-2 in our patients, a 6-year-old boy in Guangzhou, Guangdong Province, China, was reported to be under more than 34 days of hospitalization due to prolonged presence of viral RNA in stools after showing negative in respiratory samples. Actually, this phenomenon is not rare among pediatric patients.^27^ Xiao and colleagues demonstrated that about a quarter of COVID-19 patients had SARS-CoV-2 RNA detectable in feces after viral clearance in respiratory tract.^25^ However, the researchers did not discuss whether persistent shedding of SARS-CoV-2 was more common in certain age group than the others (patient’s ages ranged from 10 months to 78 years in this study). Therefore, discharged patients might be a potential source of transmission and follow-up after hospital discharge or discontinuation of quarantine is advisable.

In face of the emerging infectious disease and limited data on pediatric patients, it is unclear what role children play in SARS-CoV-2 transmission or to what extent children are affected by the virus. More attention should be drawn to children, especially for young children who cannot handle their own excretions. Caregivers of these children should avoid direct contact with the stool and close the lid before flushing the toilet. Precautionary measures are needed when conducting aerosol-generating procedures and sewer system should be kept unobstructed. In some rural areas and underdeveloped regions with poor sanitation in China, stools are often used as natural fertilizer in agriculture. As sewage cannot be properly disinfected in these places, it might be possible that SARS-CoV-2 polluted water causes infection in wild animals which bring the virus back to humans, creating a vicious cycle. It is also of vital importance to implement strict hygiene measures after reopening of kindergartens and schools to prevent spreading of the infection among preschool and schoolchildren.

The limitations of this study should also be noted. In response to the emerging disease, only respiratory specimens were required for the detection of SARS-CoV-2 according to clinical guidelines in the early stage of COVID-19 outbreak. We therefore failed to obtain stool samples from the patients during their first few days of hospitalization and could not determine whether throat swabs and fecal samples showed positive on RT-PCR analysis simultaneously. Moreover, we did not culture the virus isolated from the feces to test the viability nor measure viral loads in the samples due to limited conditions.

Taken together, there is an urgent need to re-evaluate the current version of *Diagnosis and Treatment Plan of Corona Virus Disease 2019* (tentative seventh edition).^13^ Nucleic acid testing on fecal specimens should be added to the current criteria for hospital discharge and release of isolation. We call for multi-center studies with larger sample size to clarify the time lag between fecal and respiratory specimens negative for RNA RT-PCR analysis in infected children.

### Postscript

We reported to the local health authority immediately once we noticed the persistent shedding of SARS-CoV-2 in feces of our pediatric patients about three weeks ago. When this paper was finalized, the seventh edition of *Diagnosis and Treatment Plan of Corona Virus Disease 2019* had just been issued and indicated the possibility of fecal-oral transmission of SARS-CoV-2. Currently, capable hospitals in some parts of China (such as Shanghai, Guangdong Province, and Shandong Province) have already included negative nucleic acid testing result of fecal specimens as one of the standards for hospital discharge and release of isolation.

## Data Availability

All data generated or used during the study appear in the submitted article.

## Author Contributions

Q-SX had full access to all the data in the study and took responsibility for the integrity of and the accuracy of the data. Q-SX and Y-HX designed the study and conceptualized the paper. WN, QW, G-JL, J-NT and X-FS collected the epidemiological, clinical, laboratory and radiological data. W-JL contributed to laboratory testing. WN and G-JL summarized the data. Y-HX, WN and QW wrote the initial draft of the manuscript. All authors provided critical feedback and approved the final version.

## Acknowledgements

We thank Municipal Centre of Disease Control and Prevention of Qingdao for coordinating data collection for COVID-19 patients in Qingdao. We thank all patients and their families involved in the study. We thank all health-care workers involved in the diagnosis and treatment of patients. And we thank Prof Gary WK Wong for guidance in study design and interpretation of results.

